# Defining cost-effectiveness of mechanical circulatory support escalation among patients with acute myocardial infarction complicated by cardiogenic shock

**DOI:** 10.64898/2026.09.08.26362578

**Authors:** Abbish Kamalakkannan, Yang Yang, Riley Batchelor, Brendan Backhouse, Katherine Munro, Fiona Groen, Fumitaka Yanase, James Anstey, Andrew J. Taylor, John Dyett, Craig French, Forbes McGain, Jeffrey Lefkovits, Nicholas Cox, William Chan

## Abstract

**Background:** Temporary mechanical circulatory support (MCS) for acute myocardial infarction complicated by cardiogenic shock (AMI-CS) includes microaxial-flow pumps (mFP), venoarterial extracorporeal membrane oxygenation (VA-ECMO), and combined mFP-ECMO support. These strategies differ substantially in haemodynamic capability and resource use, but their short-term cost-effectiveness has not been compared within a common decision framework.

**Methods:** We developed a 4-strategy decision-analytic model comparing No MCS (standard care without temporary MCS), mFP, VA-ECMO, and mFP-ECMO from the perspective of a publicly funded Australian health system over 30 days. Costs were estimated from individual resource components in 2025–26 Australian dollars and benchmarked against hospital casemix data. The primary outcome was cost per additional 30-day survivor. Scenario analyses and probabilistic sensitivity analysis assessed parameter uncertainty.

**Results:** Expected per-patient costs were A$34,562 for No MCS, A$125,269 for mFP, A$133,573 for VA-ECMO, and A$177,481 for mFP-ECMO. Assigned 30-day survival probabilities were 56.1%, 74.2%, 54.3%, and 53.9%, respectively. mFP was the only non-dominated MCS strategy, with an incremental cost-effectiveness ratio of A$502,695 per additional survivor compared with No MCS. Under the assigned cross-cohort inputs, VA-ECMO and mFP-ECMO were more costly and less effective, remained strictly dominated across all prespecified scenarios, and required 30-day survival above 74.2% to escape dominance. Intensive care unit (ICU) bed-days were the largest cost component. At a willingness-to-pay threshold of A$500,000 per additional survivor, the probabilities of being optimal were 44.0% for No MCS, 54.1% for mFP, 1.7% for VA-ECMO, and 0.2% for mFP-ECMO.

**Conclusions:** Under the assigned cross-cohort inputs, escalation of MCS beyond mFP in AMI-CS increased costs without improving modelled 30-day survival. Nevertheless, VA-ECMO and mFP-ECMO remain essential for selected patients requiring greater circulatory or respiratory support.

**What Is Known:** • Acute myocardial infarction complicated by cardiogenic shock (AMI-CS) carries high short-term mortality and substantial healthcare costs. Temporary mechanical circulatory support (MCS) strategies differ markedly in clinical effectiveness, complication profiles, acquisition costs, and critical-care resource use.

• Existing economic evaluations have largely been limited to pairwise comparisons and have not assessed escalation from No MCS to multiple MCS strategies within a single cost-effectiveness framework in a public, government-funded healthcare setting.

**What the Study Adds:** • A common cost-effectiveness frontier comparing No MCS with mFP, VA-ECMO, and mFP-ECMO showed that mFP was the only non-dominated MCS strategy, with an incremental cost-effectiveness ratio of A$502,695 per additional 30-day survivor.

• VA-ECMO and mFP-ECMO would each require 30-day survival exceeding 74.2% to avoid economic dominance. VA-ECMO remained dominated even when its device acquisition cost was reduced to zero.

• ICU bed-days, rather than device acquisition alone, were the principal cost driver in every arm, identifying critical-care resource use as the factor with the greatest influence on value.

## 1 Introduction

Acute myocardial infarction complicated by cardiogenic shock (AMI-CS) is associated with substantial early mortality of up to approximately 50% despite rapid revascularisation and contemporary intensive care. In a recent Australian registry, approximately 44% of patients with AMI-CS undergoing percutaneous coronary intervention (PCI) died within 30 days [1], and outcomes have not improved materially over successive treatment eras [2].

Currently available mechanical circulatory support (MCS) devices differ substantially in their circulatory support, adverse-event profiles, initial costs, and resource requirements. The DanGer Shock randomised controlled trial (RCT) demonstrated a survival benefit at 180 days from left ventricular microaxial-flow pump (mFP) support over standard care in AMI-CS, at the cost of more serious adverse events [3]. By contrast, an individual patient-data meta-analysis of 4 RCTs found that early venoarterial extracorporeal membrane oxygenation (VA-ECMO) did not improve 30-day survival and increased bleeding and peripheral vascular complications in AMI-CS [4]. The clinical value of MCS therefore cannot be inferred from the modality or intensity of support alone.

The economic evidence of MCS cost-effectiveness is similarly not well defined across different systems of care. Previous evaluations have been pairwise: mFP against VA-ECMO over a lifetime horizon in Australia [5] and Italy [6], and mFP-ECMO against guideline-directed medical therapy in Germany [7]. These comparisons do not represent the full range of strategies available to hospitals deciding whether, and how far, to escalate support. We therefore developed a 30-day decision model comparing No MCS, mFP, VA-ECMO, and mFP-ECMO to identify which strategies lay on the efficiency frontier, estimate the cost per additional 30-day survivor, and determine the survival and MCS device-price thresholds at which the ordering would change.

## 2 Methods

### 2.1 Study Design, Perspective, and Reporting

We developed a decision-analytic model comparing 4 temporary MCS strategies for AMI-CS from the perspective of a metropolitan public (government) funded health service in Victoria, Australia, over a 30-day time horizon. All costs were valued in 2025–26 Australian dollars (A$). Discounting was not applied because all costs and outcomes occurred within 30 days. The study was reported in accordance with the Consolidated Health Economic Evaluation Reporting Standards 2022 statement [8].

The completed checklist, model equations, and detailed input derivations are provided in the Supplemental Material.

#### Data Availability

The model code, health economic analysis plan, parameter table, and analysis outputs are available from the corresponding author on reasonable request. All clinical inputs were obtained from the cited published sources. The hospital casemix extract used for top-down validation is governed by the health service and cannot be shared; it did not inform any base-case input.

#### Independent Data Access and Analysis

Abbish Kamalakkannan had full access to all study data and takes responsibility for the integrity of the data and the accuracy of the data analysis.

### 2.2 Decision Model, Population, and Comparators

The modelled population comprised adults undergoing primary PCI for AMI-CS who were considered for temporary MCS. The 4 strategies were No MCS, mFP, peripheral VA-ECMO, and mFP-ECMO. The No MCS arm represented Victorian PCI-based usual care, comprising PCI-based treatment and standard care without temporary MCS. The percutaneous left ventricular mFP was the Impella CP (Abiomed, Johnson & Johnson MedTech, Danvers, MA, USA), the pump in use at the study health service over the costing period. Peripheral VA-ECMO provides both circulatory and respiratory support. mFP-ECMO, also termed ECpella, combined VA-ECMO with mFP to provide left ventricular unloading with extracorporeal support.

A decision tree assigned patients in each arm to survival or death at 30 days (Figure S1). Because the source cohorts did not report resource use separately for survivors and non-survivors, the same expected hospital cost was applied to both terminal outcomes. Each model arm was assigned a 30-day survival probability from a separate published cohort (Table 1).

**Table 1:**
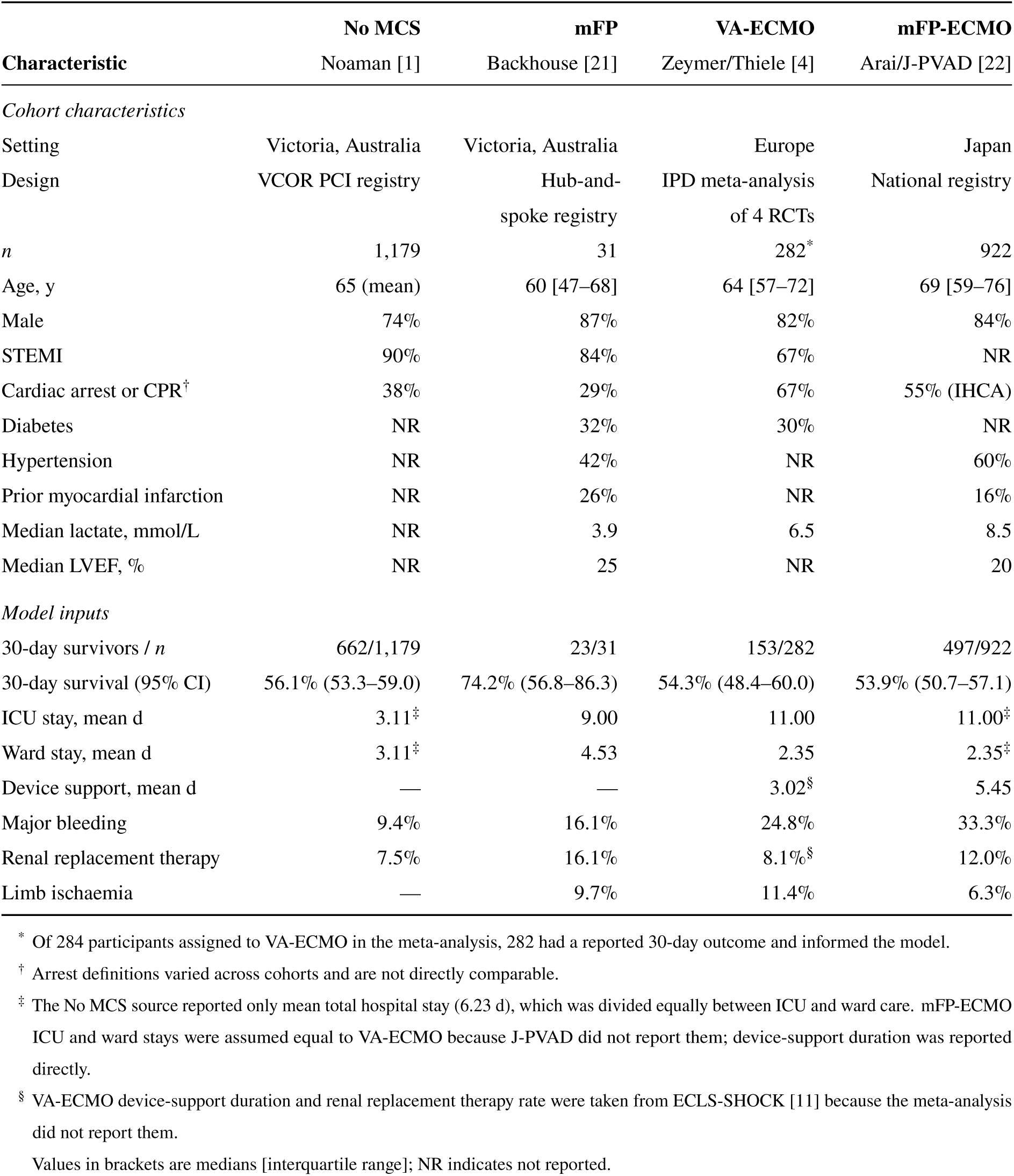
Source cohorts and clinical model inputs. Cohort characteristics describe the populations from which each strategy’s outcomes were drawn; model inputs are the values actually entered into the decision tree.

| Characteristic | No MCS<br>Noaman [1] | mFP<br>Backhouse [21] | VA-ECMO<br>Zeymer/Thiele [4] | mFP-ECMO<br>Arai/J-PVAD [22] |
| --- | --- | --- | --- | --- |
| *Cohort characteristics* |  |  |  |  |
| Setting | Victoria, Australia | Victoria, Australia | Europe | Japan |
| Design | VCOR PCI registry | Hub-and-spoke registry | IPD meta-analysis of 4 RCTs | National registry |
| *n* | 1,179 | 31 | 282* | 922 |
| Age, y | 65 (mean) | 60 [47–68] | 64 [57–72] | 69 [59–76] |
| Male | 74% | 87% | 82% | 84% |
| STEMI | 90% | 84% | 67% | NR |
| Cardiac arrest or CPR<sup>†</sup> | 38% | 29% | 67% | 55% (IHCA) |
| Diabetes | NR | 32% | 30% | NR |
| Hypertension | NR | 42% | NR | 60% |
| Prior myocardial infarction | NR | 26% | NR | 16% |
| Median lactate, mmol/L | NR | 3.9 | 6.5 | 8.5 |
| Median LVEF, % | NR | 25 | NR | 20 |
| *Model inputs* |  |  |  |  |
| 30-day survivors / *n* | 662/1,179 | 23/31 | 153/282 | 497/922 |
| 30-day survival (95% CI) | 56.1% (53.3–59.0) | 74.2% (56.8–86.3) | 54.3% (48.4–60.0) | 53.9% (50.7–57.1) |
| ICU stay, mean d | 3.11<sup>‡</sup> | 9.00 | 11.00 | 11.00<sup>‡</sup> |
| Ward stay, mean d | 3.11<sup>‡</sup> | 4.53 | 2.35 | 2.35<sup>‡</sup> |
| Device support, mean d | — | — | 3.02<sup>§</sup> | 5.45 |
| Major bleeding | 9.4% | 16.1% | 24.8% | 33.3% |
| Renal replacement therapy | 7.5% | 16.1% | 8.1%<sup>§</sup> | 12.0% |
| Limb ischaemia | — | 9.7% | 11.4% | 6.3% |

\* Of 284 participants assigned to VA-ECMO in the meta-analysis, 282 had a reported 30-day outcome and informed the model.
<sup>†</sup> Arrest definitions varied across cohorts and are not directly comparable.
<sup>‡</sup> The No MCS source reported only mean total hospital stay (6.23 d), which was divided equally between ICU and ward care. mFP-ECMO ICU and ward stays were assumed equal to VA-ECMO because J-PVAD did not report them; device-support duration was reported directly.
<sup>§</sup> VA-ECMO device-support duration and renal replacement therapy rate were taken from ECLS-SHOCK [11] because the meta-analysis did not report them.
Values in brackets are medians [interquartile range]; NR indicates not reported.

The primary outcome was the incremental cost-effectiveness ratio (ICER) per additional 30-day survivor. We also estimated the cost per 30-day survivor within each strategy. A strategy was considered strictly dominated if it cost more and produced fewer survivors than an alternative; extended dominance was assessed during construction of the efficiency frontier.

A 30-day horizon was selected because differences in follow-up between the source cohorts precluded credible extrapolation of longer-term survival and utility. The outcome was therefore expressed as a 30-day survivor rather than a quality-adjusted life-year (QALY). Unlike the QALY, this outcome has no generally accepted willingness-to-pay threshold.

### 2.3 Clinical and Cost Inputs

Table 1 summarises the 30-day survival, intensive care unit (ICU) and ward length of stay, device-support duration, and complication rates assigned to each arm. Where necessary, means and standard deviations were estimated from reported medians and interquartile ranges using established methods [9, 10]; values unavailable from a source were imputed as described in Table 1.

Survival, length of stay, and complication rates for VA-ECMO were obtained from the individual patient-data meta-analysis [4]. Renal replacement therapy and ECMO duration were retained from ECLS-SHOCK because the meta-analysis did not report these measures by treatment arm [11]. Major bleeding, renal replacement therapy, and limb ischaemia were costed separately because both arm-specific event rates and identifiable resource use were available. Stroke and sepsis were not assigned separate costs given that their principal resource consequence, prolonged critical care, was already captured through the modelled bed-days. Escalation from mFP to VA-ECMO was represented by the mFP-ECMO arm. The mFP arm did not include a rescue-ECMO cost in the base case.

Costs were estimated from the quantities and unit prices of individual resource components. These included ICU and ward bed-days, device or circuit acquisition, insertion and removal procedures, ECMO maintenance, enhanced ECMO nursing, acute PCI, and complication-related resource use. Bed-day costs were obtained from health-service finance data, while hardware prices were based on supplier quotations. Procedure costs were drawn from the Australian Medicare Benefits Schedule (MBS) [12]. Published Australian sources informed the costs of ICU diagnostic services [13], ECMO maintenance and decannulation [14], renal replacement therapy [15], and limb revascularisation [16]. The enhanced ECMO nursing item is the incremental cost of the additional nurse rostered to a patient on an active extracorporeal circuit, over and above the one-to-one ratio already priced into the ICU bed-day. It is charged over the days of device support only; its scope is site-specific and is tested in the structural scenarios (Supplemental Material). Blood products were valued using the National Blood Authority price list, and historical estimates were inflated to 2025–26 values using Reserve Bank of Australia indices [17]. Table S1 reports every unit cost, its sensitivity range, assigned probabilistic distribution, and source.

Ranges were bounded by external evidence where it existed and by a symmetric ±25% swing where it did not. Costs were given gamma distributions; schedule fees and quoted prices were held fixed. Table S1 explains how the ranges, the distributions, and the coefficients of variation were derived.

### 2.4 Analysis

Expected cost was defined as the mean cost per patient across survivors and non-survivors rather than the cost of an individual admission (Supplemental Material). Incremental analysis ordered the strategies by increasing effectiveness after strictly and extendedly dominated alternatives had been removed.

Willingness to pay was defined as the amount a decision maker would pay for one additional 30-day survivor. At each willingness-to-pay value, net monetary benefit was calculated as the survival probability multiplied by willingness to pay, minus expected cost. The strategy with the greatest net monetary benefit was considered optimal. This framework allowed all 4 strategies to be evaluated simultaneously without relying on potentially unstable pairwise ICERs. Values from A$0 to A$1,000,000 per additional survivor were examined in A$10,000 increments.

### 2.5 Uncertainty, Scenario, and Structural Analyses

In the one-way sensitivity analysis, each input was varied separately across the range reported in Table S1. Outcomes included the cost per 30-day survivor for each strategy and the pairwise ICER for mFP versus VA-ECMO.

Prespecified resource and costing scenarios are set out in Table S2. Two further scenarios examined structural assumptions in the ECMO arms: the enhanced ECMO nursing premium extended from the period of device support to the entire ICU stay, and mFP-ECMO support duration varied across its reported interquartile range.

In the probabilistic analysis, the model was run 10,000 times, with every uncertain input redrawn from the distribution assigned to it in Table S1. Unit costs shared between strategies, such as the intensive care bed-day rate, were drawn once per run and applied to every arm; clinical quantities were drawn separately for each arm because they came from different source cohorts. We reported mean outcomes across the 10,000 runs with 95% uncertainty intervals defined by the 2.5th and 97.5th percentiles, the proportion of runs in which each strategy had the greatest net monetary benefit, and the proportion in which one strategy was both more costly and less effective than another. The model was also checked against 10 boundary and identity conditions with analytically known answers, all of which were satisfied.

### 2.6 Threshold, Value-of-Information, and Budget-Impact Analyses

Threshold analyses examined the 30-day survival at which each ECMO strategy would cease to be dominated, the minimum mFP survival required to remain non-dominated, and the mFP device price at which its total arm cost would equal that of each ECMO strategy. Additional analyses removed ECMO hardware costs entirely, varied rescue-ECMO escalation from 0% to 50%, and varied mFP and VA-ECMO survival jointly while holding costs constant.

Expected value of perfect information was estimated across the willingness-to-pay range and scaled to the annual state-wide cohort. Its derivation is described in the Supplemental Material.

Per-patient results were scaled to 295 AMI-CS cases per year for Victoria, Australia, based on the Victorian Cardiac Outcomes Registry PCI cohort [1].

### 2.7 Cost-Model Validation and Statistical Analysis

The bottom-up mFP estimate was compared for face validity with a top-down casemix extract of consecutive costed mFP admissions at the study health service. Of 42 episodes, one was excluded for an implausible recorded cost with no assigned diagnosis-related group. This extract was used only for external validation and did not inform any base-case input. Analyses were performed in Python. Wilson intervals were used for the observed survival proportions in Table 1, while model uncertainty intervals were derived from the percentiles of the specified parameter distributions.

### 2.8 Ethics

This economic evaluation used published clinical inputs, local health-service financial data, supplier quotations, and aggregate clinical data approved by the Alfred Human Research Ethics Committee (Project Number 211/21). A hospital casemix extract governed by the health service was used solely for external cost-model validation and did not inform any clinical-effectiveness or base-case cost input. No additional study-specific ethics approval was required.

## 3 Results

### 3.1 Base-Case Cost-Effectiveness

Across the 4 strategies, expected cost ranged from A$34,562 per patient for No MCS to A$177,481 for mFP-ECMO, while assigned 30-day survival probabilities ranged from 53.9% to 74.2% (Table 2, Panel A). Compared with No MCS, mFP increased expected cost by A$90,708 and modelled survival by 18.0 percentage points, producing an ICER of A$502,695 per additional 30-day survivor. By contrast, mFP cost A$8,304 less than VA-ECMO and had a 19.9-percentage-point higher survival estimate.

**Table 2:**
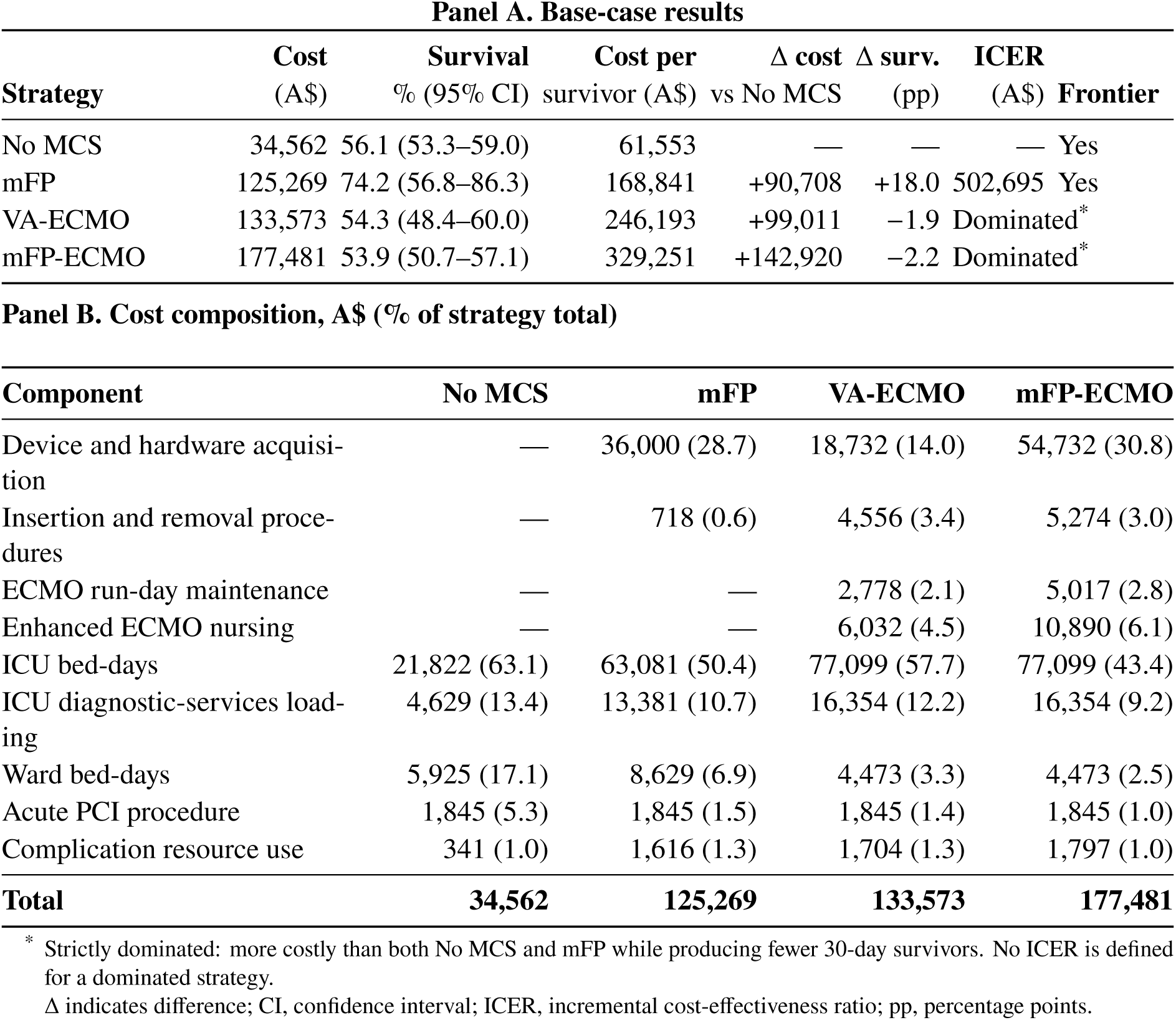
Base-case cost-effectiveness (Panel A) and per-patient cost composition (Panel B).

Under the assigned cross-cohort survival estimates, VA-ECMO and mFP-ECMO were both more costly and had lower assigned survival than No MCS and mFP, and were therefore strictly dominated. The efficiency frontier comprised only No MCS and mFP.

### 3.2 Cost Composition and External Validation

ICU bed-days were the largest individual cost component in every strategy, ranging from 43.4% of mFP-ECMO cost to 63.1% of No MCS cost (Table 2, Panel B). Device and hardware acquisition represented 28.7% of mFP cost and 30.8% of mFP-ECMO cost, whereas the enhanced ECMO nursing premium, charged over the device support run, contributed 4.5% of VA-ECMO cost and 6.1% of mFP-ECMO cost. The top-down validation extract comprised 41 mFP admissions, for which the median cost was A$139,947 (interquartile range A$112,718–A$198,923). The bottom-up estimate of A$125,269 was 10.5% below the median. An independent check on the No MCS arm from the same casemix system gave approximately A$39,000 against the modelled A$34,562 [18]; the derivation is given in the Supplemental Material.

### 3.3 One-Way and Scenario Analyses

Across all 4 strategies, cost per survivor was most sensitive to 30-day survival and ICU resource use (Figure 1). Device acquisition cost exerted less influence than either factor in every strategy that included a device. The comparison between mFP and VA-ECMO was driven primarily by survival and ICU length of stay, and variation across plausible bounds could change strict dominance into a more-costly, more-effective trade-off.

**Figure 1:**
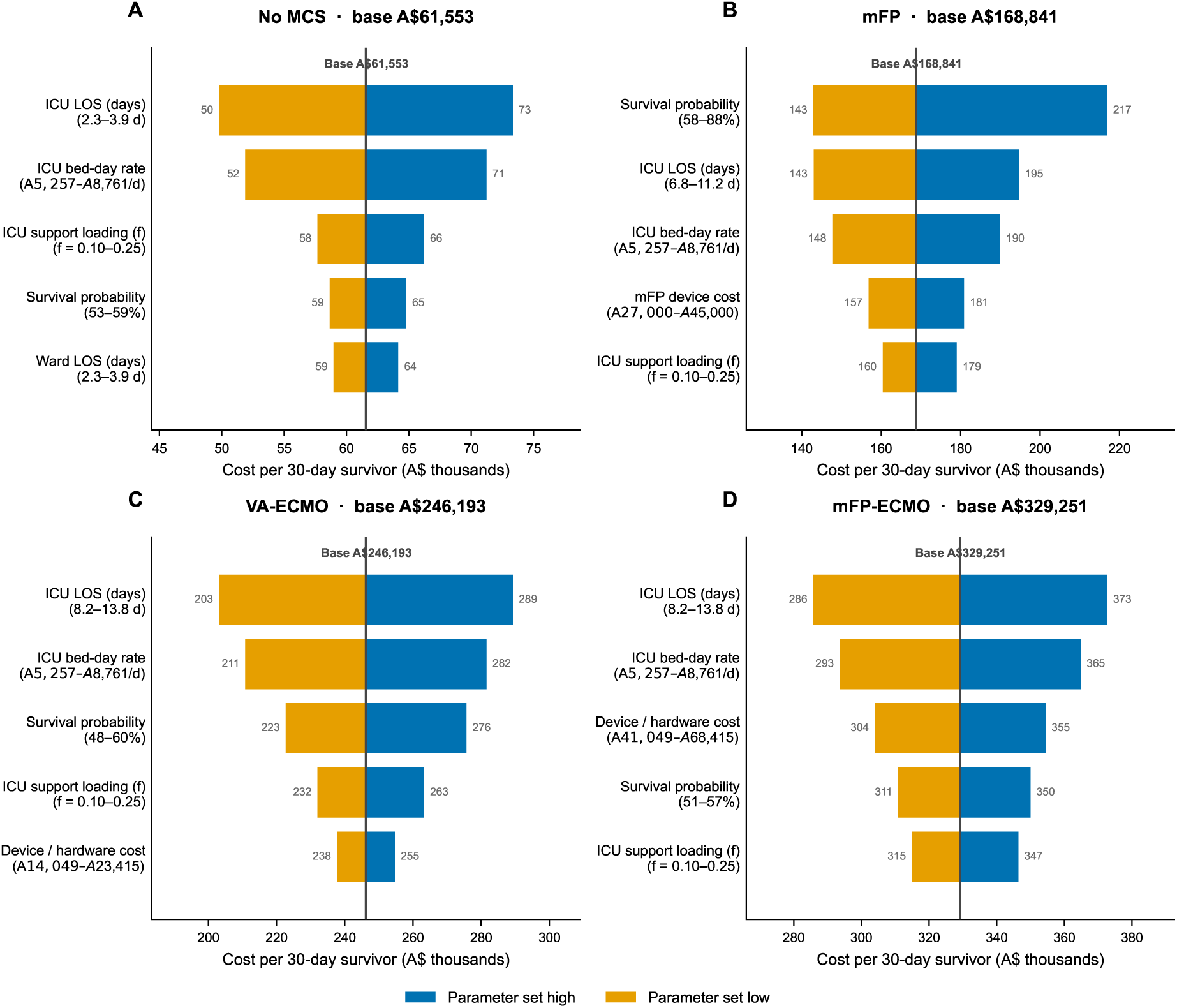
One-way sensitivity analysis of cost per 30-day survivor. Each panel shows the 5 most influential inputs for one strategy. Bar labels give A$ thousands per 30-day survivor at the lower and upper bounds. Colour identifies the bound. Table S1 gives the input ranges. A$ indicates Australian dollars; CCU, coronary care unit; d, day; *f* , diagnostic-services share of a full ICU day; ICU, intensive care unit; LOS, length of stay; MCS, mechanical circulatory support; mFP, microaxial-flow pump; VA-ECMO, venoarterial extracorporeal membrane oxygenation.

The frontier was unchanged in every prespecified scenario, resource, costing, and structural alike (Table S2). Across the resource and costing scenarios, the ICER for mFP versus No MCS ranged from A$454,193 to A$530,410. It increased to A$584,038 when the top-down median of A$139,947 replaced the bottom-up mFP cost (Table S4).

Extending the enhanced nursing premium from the device support run to the whole ICU stay increased the expected cost of VA-ECMO to A$149,541 and that of mFP-ECMO to A$188,591, widening rather than narrowing the gap to mFP. Varying mFP-ECMO support duration across its interquartile range changed its expected cost from A$168,293 to A$187,282.

### 3.4 Probabilistic Analysis

Although uncertainty intervals overlapped substantially across strategies, the probabilistic mean costs retained the base-case ranking (Table 3). mFP was more effective than No MCS in 97.9% of iterations, with the comparison representing a more-costly, more-effective trade-off in 91.0% (Figure 2).

**Figure 2:**
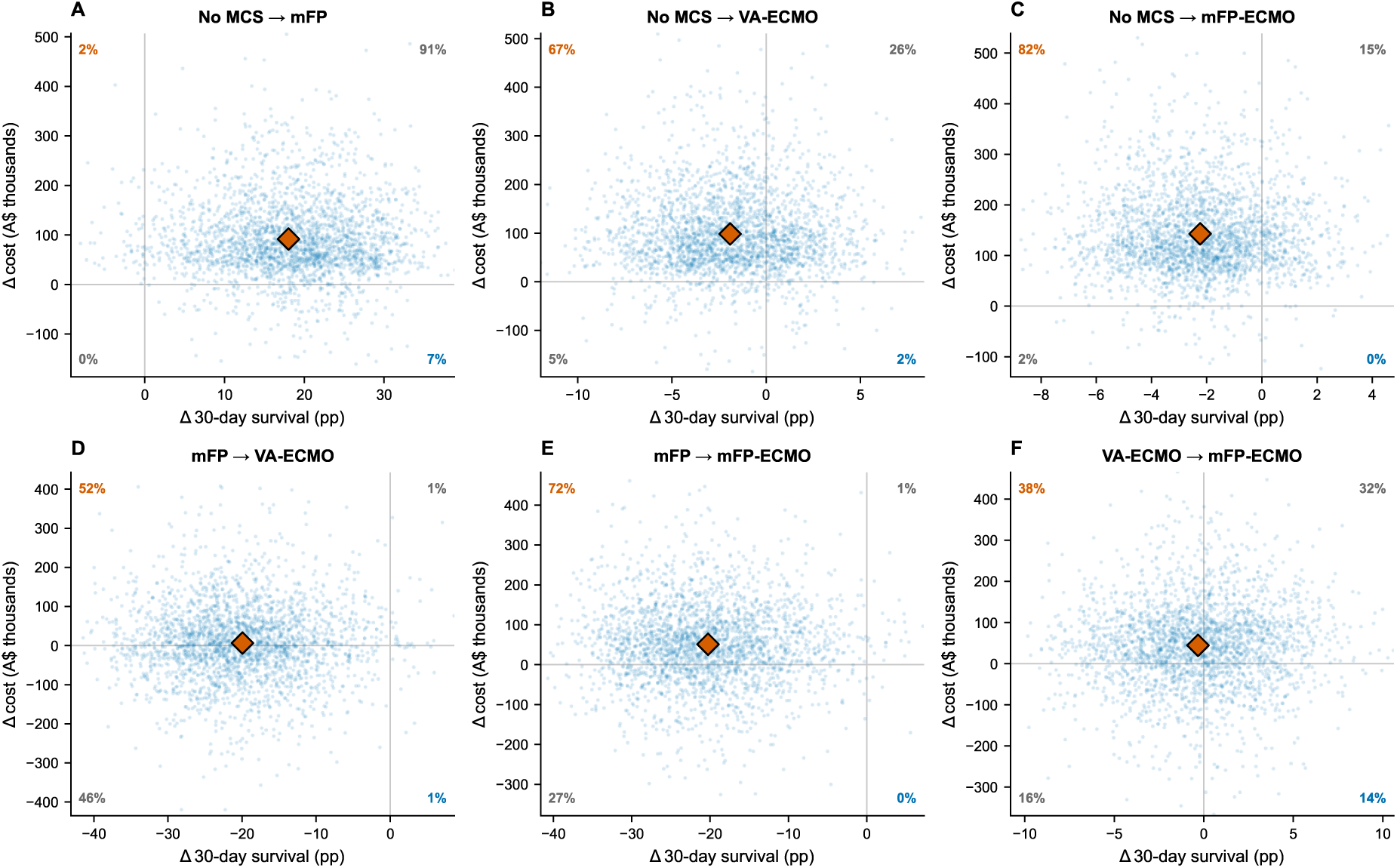
Probabilistic cost-effectiveness planes for each pairwise escalation. Each panel shows the difference in cost and 30-day survival across 10,000 iterations. The diamond marks the mean. Corner values give the percentage of iterations in each quadrant and sum to 100%. Orange (upper left) shows the probability that the more intensive strategy is dominated. Blue (lower right) shows the probability that it is dominant. Panels A–F present the labelled comparisons from left to right, top to bottom; arrows indicate the direction from the reference strategy to the comparator. Δ indicates difference; A$, Australian dollars; ECMO, extracorporeal membrane oxygenation; MCS, mechanical circulatory support; mFP, microaxial-flow pump; pp, percentage points; VA-ECMO, venoarterial ECMO.

**Table 3:** Probabilistic sensitivity analysis, 10,000 iterations: expected cost, expected survival, and the probability that each strategy is optimal.

| Strategy | Mean cost, A\$<br>(95% UI) | Mean survival,<br>% (95% UI) | Probability optimal (%) | | | |
| --- | --- | --- | --- | --- | --- | --- |
| | | | A\$<br>100,000 | A\$<br>300,000 | A\$<br>500,000 | A\$<br>700,000 |
| No MCS | 34,385 (4,841–<br>144,710) | 56.2 (53.3–<br>59.0) | 85.4 | 64.5 | 44.0 | 30.9 |
| mFP | 126,229 (49,163–<br>321,017) | 74.2 (57.1–<br>88.1) | 10.0 | 32.8 | 54.1 | 67.8 |
| VA-ECMO | 132,747 (43,551–<br>334,955) | 54.2 (48.3–<br>60.0) | 4.2 | 2.5 | 1.7 | 1.2 |
| mFP-ECMO | 177,331 (83,806–<br>374,925) | 53.9 (50.7–<br>57.1) | 0.5 | 0.3 | 0.2 | 0.1 |

A strategy is optimal when it has the greatest net monetary benefit. Probabilities sum to 100% within each column, subject to rounding. UI indicates the 2.5th to 97.5th percentile of the simulated distribution.

Relative to mFP, VA-ECMO and mFP-ECMO were strictly dominated in 51.9% and 71.7% of iterations, respectively, and were more effective in only 1.6% and 1.1% (Figure 2). The comparison between the 2 ECMO strategies was less clearly resolved: mFP-ECMO was more effective than VA-ECMO in 46.1% of iterations.

### 3.5 Willingness to Pay and Value of Information

Across the willingness-to-pay range, the optimal strategy changed only between No MCS and mFP (Figure 3, Table S5). At A$500,000 per additional survivor, the probabilities of being optimal were 44.0% for No MCS, 54.1% for mFP, 1.7% for VA-ECMO, and 0.2% for mFP-ECMO; the values at A$100,000 and A$700,000 are given in Table 3.

**Figure 3:**
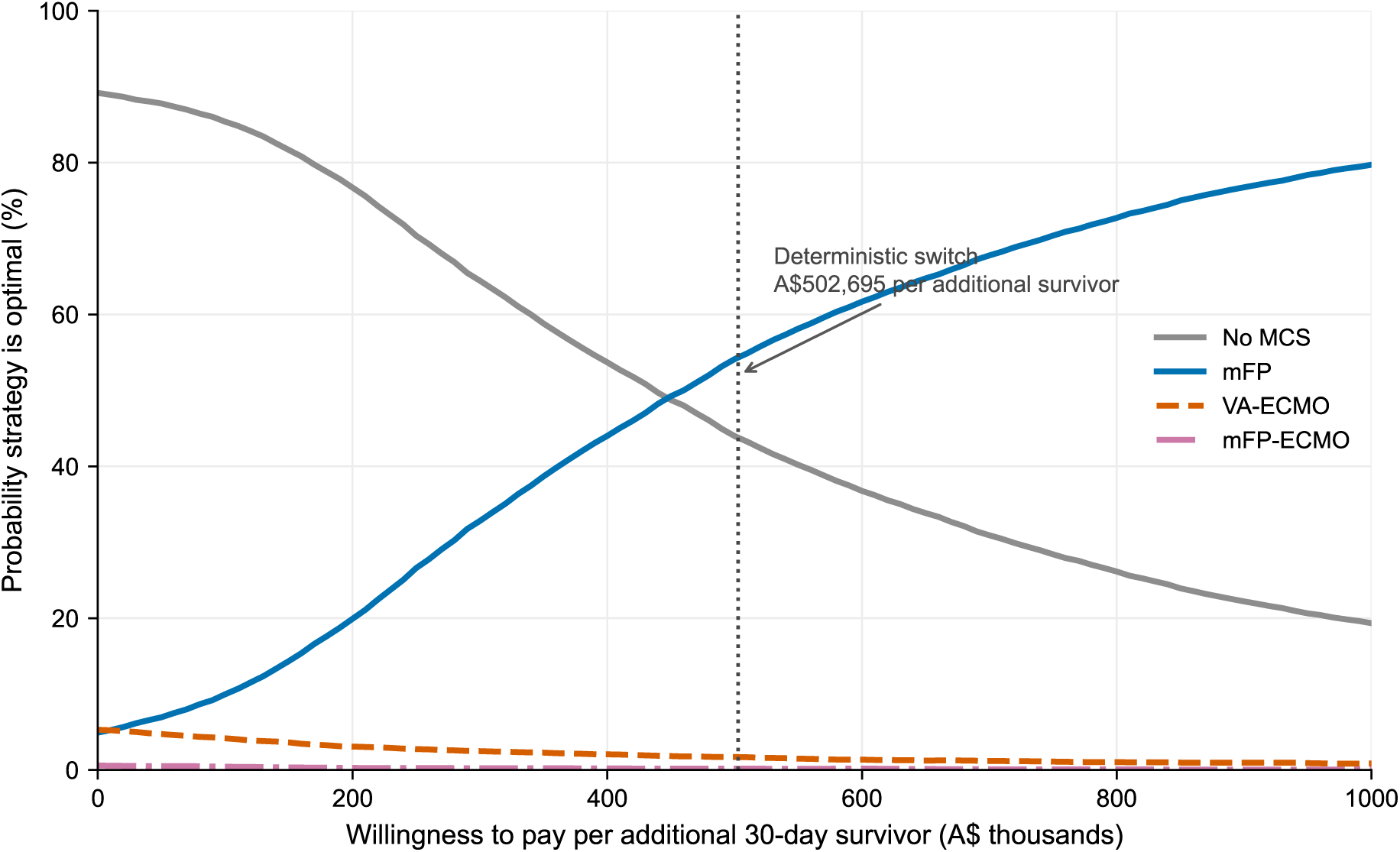
Cost-effectiveness acceptability curves across 10,000 probabilistic iterations. Each line gives the probability that a strategy has the greatest net monetary benefit at the corresponding willingness to pay per additional 30-day survivor. The dotted line marks the deterministic switch point of A$502,695 per additional 30-day survivor. A$ indicates Australian dollars; ECMO, extracorporeal membrane oxygenation; MCS, mechanical circulatory support; mFP, microaxial-flow pump; VA-ECMO, venoarterial ECMO.

mFP first became more likely to be optimal than No MCS at A$450,000 per additional survivor, close to the deterministic net-benefit switch point of A$502,695 (Table 4). Neither ECMO strategy had a probability of being optimal greater than 5.3% at any value examined. Expected value of perfect information reached a maximum of A$34,785 per patient at a willingness to pay of A$510,000, equivalent to A$10.26 million annually when applied to 295 patients.

**Table 4:** Threshold and willingness-to-pay analyses.

| Analysis | Base value | Threshold | Interpretation |
| --- | --- | --- | --- |
| VA-ECMO survival to escape dominance | 54.3% | > 74.2% | 19.9 points above the assigned value, and equal to the mFP arm |
| mFP-ECMO survival to escape dominance | 53.9% | > 74.2% | 20.3 points above the assigned value |
| mFP survival to remain non-dominated | 74.2% | > 56.1% | Must exceed No MCS survival |
| mFP device price for cost parity with VA-ECMO | A\$36,000 | A\$44,304 | Costs equal at this price; mFP remains more effective at any price |
| mFP device price for cost parity with mFP-ECMO | A\$36,000 | A\$88,212 | Costs equal at this price |
| ECMO hardware removed (service owns hardware) | A\$133,573 | A\$114,841 | VA-ECMO remains dominated by No MCS at zero hardware cost |
| Rescue-ECMO escalation at which the mFP arm costs as much as VA-ECMO | 0% (base case) | 15.9% | 0.2 points below the 16.1% observed rate; at the observed rate the ICER is A\$670,696 (Table S3) |
| Willingness-to-pay switch point | — | A\$502,695 | No MCS optimal below, mFP above; neither ECMO strategy was deterministically optimal at any value tested |

Costs are in 2025–26 Australian dollars; willingness to pay is expressed per additional 30-day survivor.

### 3.6 Threshold Analyses

VA-ECMO and mFP-ECMO each required 30-day survival above 74.2% to escape strict dominance, substantially higher than their assigned values of 54.3% and 53.9% (Table 4). The mFP device price could increase from A$36,000 to A$44,304 before the mFP arm reached cost parity with VA-ECMO, and to A$88,212 before it reached parity with mFP-ECMO. Removing VA-ECMO hardware costs entirely reduced its expected cost to A$114,841, but the strategy remained dominated by No MCS.

Incorporating rescue-ECMO escalation into the mFP arm increased its ICER in proportion to the assumed escalation rate. At the 16.1% rate reported in the Victorian series, the ICER reached A$670,696 per additional survivor. The mFP arm reached cost parity with VA-ECMO at an escalation rate of 15.9%, close to the observed rate, although its survival advantage persisted and VA-ECMO remained dominated throughout (Table S3). When mFP and VA-ECMO survival were varied jointly at fixed costs, VA-ECMO became the highest-survival non-dominated strategy only when its modelled survival exceeded that of mFP.

### 3.7 Budget Impact

At an estimated annual volume of 295 cases in Victoria, Australia, expected total expenditure was A$10.20 million with No MCS, A$36.95 million with mFP, A$39.40 million with VA-ECMO, and A$52.36 million with mFP-ECMO, so full adoption of mFP increased annual expenditure by A$26.76 million relative to No MCS for 53.2 additional 30-day survivors.

## 4 Discussion

Within this 30-day cross-cohort framework, mFP was the only MCS strategy on the efficiency frontier. VA-ECMO and mFP-ECMO were more costly and were associated with lower assigned survival in their respective source cohorts. Both remained strictly dominated across every prespecified resource, costing, and structural scenario. Because the separation between arms arose primarily from the assigned survival estimates rather than device costs, even substantial reductions in hardware cost did not restore either ECMO strategy to the frontier.

These findings concern the economics and financial implications of adopting each strategy when MCS is considered or required for the treatment of AMI-CS. The present data should not be interpreted as a causal ranking of the devices themselves. The analysis provides a health-service perspective on the trade-offs encountered when implementing an MCS program for AMI-CS. Each arm inherited its outcome from a different source cohort, with important differences in case mix, study design, and treatment era. The direction of the modelled survival difference is nevertheless consistent with the available observational head-to-head evidence: a systematic review of 6 cohort studies including 7,093 patients with AMI-CS reported lower in-hospital mortality with mFP than with VA-ECMO (risk ratio 0.89, 95% CI 0.83 to 0.96; 0.72, 95% CI 0.59 to 0.86 in propensity-matched analyses), in the absence of any randomised comparison [19]. VA-ECMO and mFP-ECMO also provide biventricular and respiratory support that a left ventricular microaxial-flow pump cannot. Their clinical role in appropriately selected patients was not assessed in the current study.

Previous economic evaluations have considered these strategies only in pairs. An Australian lifetime analysis using propensity-matched effectiveness estimates found mFP support to be cost-saving relative to VA-ECMO by A$30,157 per patient, alongside a gain in QALYs [5]. An Italian lifetime Markov analysis reached a similar conclusion from a national health-service perspective demonstrating mFP support to be dominant, with costs of €50,303 versus €76,795 and outcomes of 0.905 versus 0.784 QALYs [6]. A German analysis comparing mFP-ECMO with guideline-directed medical therapy, from a statutory health-insurance perspective, instead reported an ICER of €105,263 per QALY in an unselected population [7]. The present analysis differs from these studies in its 30-day horizon and its use of a 30-day survivor rather than a QALY. The direction of the acute cost difference agreed with the earlier Australian analysis, but its magnitude did not: mFP cost A$8,304 less than VA-ECMO here, against A$30,157 over a lifetime horizon [5], so the agreement extends only to the sign of the difference.

External estimates support the cost model. The modelled VA-ECMO cost of A$133,573 fell between two Australian benchmarks: below the Australian and New Zealand EXCEL registry casemix mean of A$215,128 in 2022 values [20], and above a whole-care-cycle estimate derived through time-driven activity-based costing [14]. This ordering is expected, given casemix costing includes system overheads that our model excluded, whereas a micro-costed care cycle captures a narrower set of activities than our model included. Neither benchmark is specific to AMI-CS, so both confirm that the modelled cost is of a plausible magnitude rather than validating its individual components. The same pattern held for mFP, where the bottom-up estimate of A$125,269 was 10.5% below the median of the top-down casemix extract at the study health service (Table S4).

Although device acquisition is the most visible expense associated with MCS, ICU bed-days were the largest cost component in every arm. This remained true for mFP-ECMO despite the acquisition of 2 devices for each patient. After the diagnostic-services loading and enhanced nursing premium were included, critical care accounted for 74.5% of the expected VA-ECMO cost. Removing VA-ECMO hardware costs did not return the strategy to the frontier, whereas the ICU diagnostic-services assumption alone changed the ICER by A$76,000. Programs seeking to improve the value of temporary MCS may therefore achieve more by examining ICU bed duration, weaning protocols, and enhanced nursing requirements than by focusing exclusively on device prices.

Device selection must remain clinically driven, since VA-ECMO and mFP-ECMO may be necessary when biventricular or respiratory support is required. Dominance within a cross-cohort model is not evidence that these technologies should be withheld when clinically indicated; rather, it argues against their routine use in patients who do not require their additional support.

The price thresholds may nevertheless inform procurement. The annual expected value of perfect information of A$10.26 million is an upper bound on the value of eliminating current parameter uncertainty rather than a proposed research budget, and it supports further prospective comparative research in which survival and resource use are collected concurrently.

The analysis has several strengths. All 4 strategies were compared on a common frontier rather than in pairs, costs were built from local and national resource components rather than overseas reimbursement tariffs, and the model was verified against 10 boundary and identity conditions and against an independent top-down casemix extract.

Because the ICER is expressed per additional 30-day survivor, it cannot be compared directly with a cost-per-QALY threshold. Dividing the ICER by a cost-per-QALY threshold gives a break-even figure: each additional survivor would need to accrue about 10.1 discounted QALYs for mFP to be cost-effective at A$50,000 per QALY, and 5.0 at A$100,000. The model assigns no post-discharge survival or utility, so this is an arithmetic translation rather than a result.

Although the base-case ordering persisted in every prespecified scenario, the probabilistic analysis appropriately showed that it was not reproduced in every individual iteration.

### 4.1 Limitations

The findings should be interpreted in light of several limitations. First, the 30-day horizon excludes subsequent survival, readmission, rehabilitation, and quality-of-life outcomes. Extending the analysis to 6 or 12 months would require arm-specific post-30-day survival, costs, and utilities, which are not currently available.

Second, no published trial has directly compared all 4 MCS strategies. The clinical inputs were therefore drawn from separate heterogeneous randomised and non-randomised cohorts that differed in country, study design, case mix, and treatment era, and the mFP survival estimate came from a small series. Its 95% confidence interval extends down to 56.8%, close to the 56.1% at or below which mFP would itself be dominated by No MCS, and the probabilistic analysis reflects this: mFP was optimal in 54.1% of iterations at A$500,000 per additional survivor. The model estimates the expected costs and outcomes associated with adopting each strategy as the default approach but does not estimate relative treatment effects. Differences between arms may reflect differences between the source populations rather than the effects of the support strategies themselves. The No MCS effectiveness cohort included some intra-aortic balloon pump and VA-ECMO use, whereas the No MCS cost model excluded temporary MCS acquisition. This structural mismatch may bias the comparison. In addition, the source cohorts did not report harmonised Society for Cardiovascular Angiography and Interventions (SCAI) shock stages, leaving substantial potential for residual confounding by disease severity.

Third, MBS fees may differ from the true resource cost incurred by a health service. No separate fee was identified for routine percutaneous mFP removal, so the surgical removal fee was included only in sensitivity analysis. Several other cost inputs were derived from a single site or from published literature and may not generalise to every health service, including the local enhanced nursing premium, whose scope is tested in scenario S1.

## 5 Conclusions

Under the assigned cross-cohort inputs in this 30-day model, mFP was the only MCS strategy on the efficiency frontier. VA-ECMO and mFP-ECMO were more costly and had lower assigned survival in their respective source cohorts, while plausible reductions in MCS device cost did not alter the ordering. These findings describe the short-term economic and clinical trade-offs associated with adopting each strategy, but they do not establish that one device causes better outcomes than another. Device selection should therefore remain guided by the support requirements of the individual patient.

## Article Information

### Author Contributions

Abbish Kamalakkannan, Yang Yang, and William Chan conceived the study and performed the analysis. All other authors contributed to the study proposal and reviewed the manuscript.

### Supplemental Material

Detailed methods; Figure S1; Tables S1–S5; CHEERS 2022 checklist.

## Non-standard Abbreviations and Acronyms

AMI-CS: acute myocardial infarction complicated by cardiogenic shock
ICER: incremental cost-effectiveness ratio
CCU: coronary care unit
ICU: intensive care unit
CI: confidence interval
IHCA: in-hospital cardiac arrest
CPR: cardiopulmonary resuscitation
IPD: individual patient data
ECMO: extracorporeal membrane oxygenation
ECLS-SHOCK: Extracorporeal Life Support in Cardiogenic Shock
J-PVAD: Japanese Registry for Percutaneous Ventricular Assist Device
LVEF: left ventricular ejection fraction
MBS: Medicare Benefits Schedule
MCS: mechanical circulatory support
mFP: microaxial-flow pump
NMB: net monetary benefit
PCI: percutaneous coronary intervention
PSA: probabilistic sensitivity analysis
QALY: quality-adjusted life-year
RCT: randomised controlled trial
SCAI: Society for Cardiovascular Angiography and Interventions
STEMI: ST-elevation myocardial infarction
UI: uncertainty interval
VA-ECMO: venoarterial extracorporeal membrane oxygenation
VCOR: Victorian Cardiac Outcomes Registry

